# Anatomical Substrates and Connectivity for Parkinson’s Disease Bradykinesia Components after STN-DBS

**DOI:** 10.1101/2023.03.02.23286704

**Authors:** Min Jae Kim, Yiwen Shi, Jasmine Lee, Yousef Salimpour, William S. Anderson, Kelly A. Mills

**Author notes:** **Corresponding Author: Kelly A. Mills, MD**, Johns Hopkins University School of Medicine, Dept. of Neurology, Meyer 6-181D, 600 N. Wolfe Street Baltimore, MD 21287. **Funding:** Kelly Mills received research funding from NIH/NINDS (5K23NS101096-01A1), Michael J. Fox Foundation, Parkinson Foundation, UCB, FDA (U01FD005942), and received honoraria from the Parkinson Study Group.

## Abstract

**Background:** Parkinsonian bradykinesia is rated using a composite scale incorporating slowed frequency of repetitive movements, decrement amplitude, and arrhythmicity. Differential localization of these movement components within basal ganglia would drive the development of more personalized network-targeted symptomatic therapies.

**Methods:** Using an optical motion sensor, amplitude and frequency of hand movements during grasping task were evaluated with subthalamic nucleus (STN)-Deep Brain Stimulation (DBS) “on” or “off” in 15 patients with Parkinson’s disease (PD). The severity of bradykinesia was assessed blindly using the MDS-UPDRS Part-III scale. Volumes of activated tissue (VAT) of each subject were estimated where changes in amplitude and frequency were mapped to identify distinct anatomical substrates of each component in the STN. VATs were used to seed a normative functional connectome to generate connectivity maps associated with amplitude and frequency changes.

**Results:** STN-DBS-induced change in amplitude was negatively correlated with change in MDS-UPDRS-III right (r = -0.65, p < 0.05) and left hand grasping scores (r = -0.63, p < 0.05). The change in frequency was negatively correlated with amplitude for both right (r = -0.63, p < 0.05) and left hand (r = -0.57, p < 0.05). The amplitude and frequency changes were represented as a spatial gradient with overlapping and non-overlapping regions spanning the dorsolateral-ventromedial axis of the STN. Whole-brain correlation maps between functional connectivity and motor changes were also inverted between amplitude and frequency changes.

**Conclusion:** DBS-associated changes in frequency and amplitude were topographically and distinctly represented both locally in STN and in whole-brain functional connectivity.

## Introduction

Deep Brain Stimulation (DBS) is a well-established treatment for refractory motor complications of Parkinson’s Disease (PD), and most often targets the subthalamic nucleus (STN) or internal pallidum. ^1, 2^ STN DBS is the most common procedure internationally, and is used to target tremor, rigidity, and bradykinesia.

To assess the baseline severity of bradykinesia and its clinical changes after therapeutic interventions, the MDS-Unified Parkinson’s Disease Rating Scale (MDS-UPDRS III) is widely adopted in clinics since it provides a reliable quantitative feature of bradykinesia severity through different tasks of both upper and lower extremities.^3^ However, bradykinesia ratings represent a composite score encompassing multiple motor features. For example, a single rating, ranging from 0 to 4, can be determined in the presence of loss of rhythm or interruptions, slowing, or amplitude decrements during the task.^3^ As such, potentially independent motor features may differentially influence the assessment of bradykinesia based on the MDS-UPDRS Part III rating.^4,^

Recent neuroimaging studies have highlighted the presence of spatially localized “sweet spots” in the STN for DBS, stimulation of which may result in superior motor improvement. ^6-8^ However, neural substrates within the STN associated with changes in individual bradykinesia motor features are not well characterized. Understanding the neural substrates representing independent motor features of bradykinesia would help elucidate the larger role STN plays in complex human motor function. Furthermore, DBS provides a unique opportunity to study the systems neuroscience of motor control by deconstructing parallel circuitry through the STN by stimulating specific sub-regions. This approach would help describe the representation of motor features of bradykinesia both within the basal ganglia and across larger cortical networks. Clinically, understanding a refined “sweet spot” for each of these motor features could aid in tailoring imaging-based DBS programming to specifically target a given patient’s symptoms and signs. It could also potentially help develop more personalized DBS therapy approaches that target the most prominent components of bradykinesia in an individual patient.

In this study, we first investigated changes in two separate motor features – amplitude and frequency – and their association with the overall MDS-UPDRS III bradykinesia rating after STN-DBS was applied. Based on these findings, we then explored how these changes in motor features were represented in both local STN stimulation sites and larger cortical network levels through functional connectivity analysis.

## Methods

### 1. Patient Recruitment

Patients diagnosed with clinically established PD^9^ who underwent bilateral STN DBS with stable stimulation parameters for at least 3 months at the Johns Hopkins Neuromodulation and Advanced Therapies Clinic were recruited for the study (Internal IRB number: IRB00270213). Patients are evaluated for advanced therapies with a multi-disciplinary approach that includes medication response testing, neuropsychological evaluation, and as-needed ancillary assessments by psychiatry or physical therapy.^10^ Patients who had dementia, language impairments, or were known to have significant discomfort when DBS was turned off were excluded from the study. Patients without sufficient postoperative CT imaging for lead reconstruction were excluded from the study.

### 2. Motor Feature Selection & Processing

We utilized the Leap Motion Controller (LMC) (Ultraleap, Mountain View, CA USA) optical tracking sensor to capture real-time hand location and assess motor features of bradykinesia. Knowledge of time-variant hand coordinates from the LMC sensor allowed us to quantify hand motion and evaluate different motor features of bradykinesia independently. LMC sensors have been adopted in previous investigations for objective motor quantification in PD. ^11-13^ Because the change in amplitude and frequency of movement is most pertinent when patients perform MDS-UPDRS III bradykinesia testing, we chose these two metrics as motor features of interest for subsequent analysis. To best represent changes in the movement of each distal fingertip captured by the LMC sensor, we chose to analyze motor metrics recorded only from the hand movement “Grasping” (GR) task administered with MDS-UPDRS-III instructions.

Amplitude and frequency motor metrics were processed and evaluated: The LMC sensor records real-time hand position by tracking individual fingertips and joints **(Figure 1A)**. From LMC recordings, we were able to reconstruct time-variant coordinates in 3D space of the (1) distal tips of four fingers (D1, D2, D3, D4) and (2) center of the palm while patients were performing the task. Raw movement waveform was captured by taking the mean euclidian distance between each point of D1, D2, D3, D4 and the center of the palm for each trial. From this waveform, amplitude and frequency were independently evaluated.

**Figure 1.**
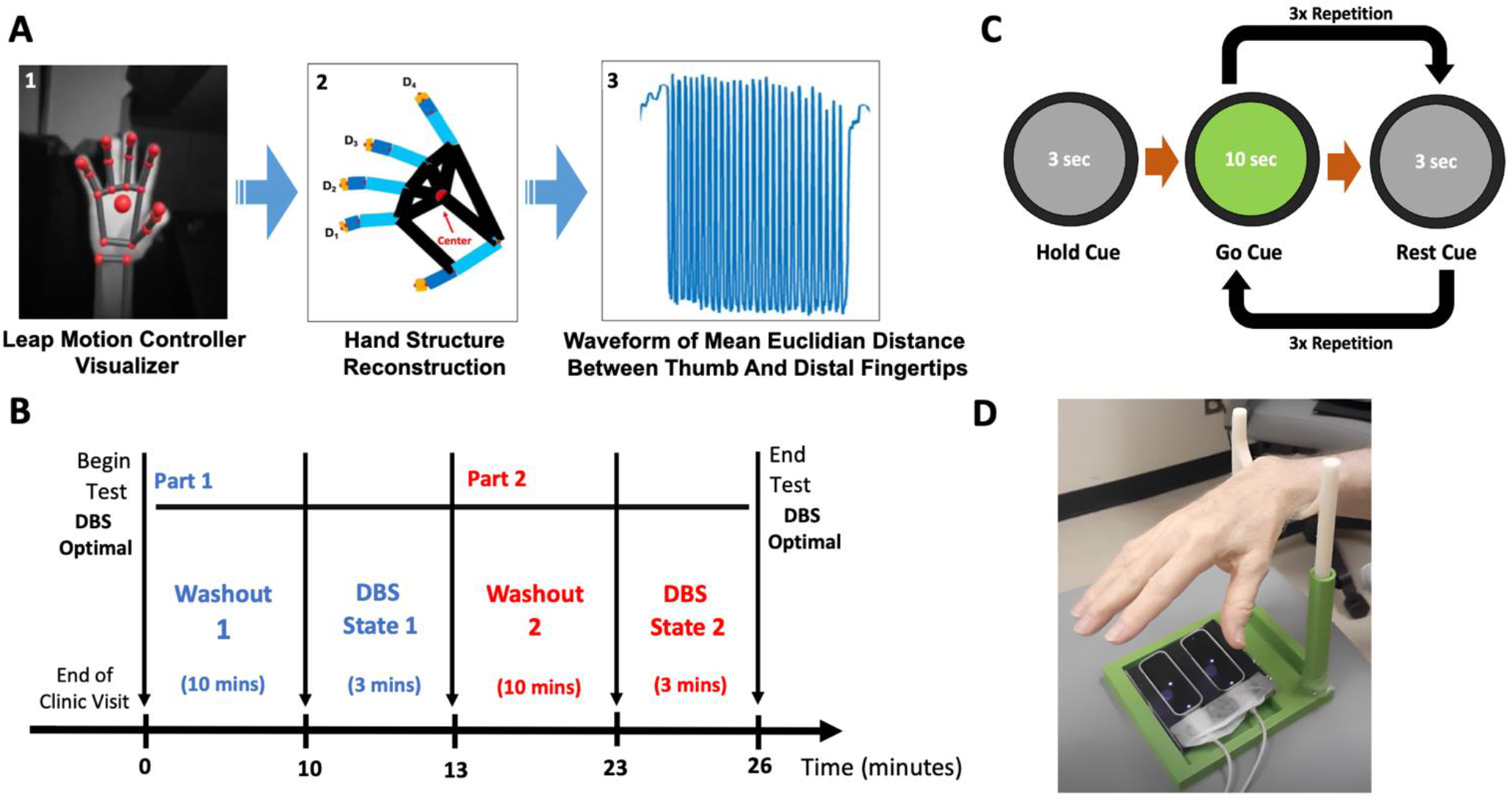
Study Design for Motor Data Collection. **(A)** Bradykinesia Motor Features Processing Flowchart: (1) Hand visualization from Leap Motion Controller (LMC) software user interface. (2) Hand coordinate extraction (D1 – D4) after LMC recording. (3) Time-variant waveform of hand movement during tasks. **(B-C)** The order of the DBS-ON and DBS-OFF was randomized and blinded to the rater, experimenter, and participant. Before each state, the study participant underwent a 10-minute washout period to remove residual DBS stimulation effects from the preceding state. **(D)** Study apparatus with optical motion sensor.

Amplitude was derived as the mean amplitude of movement waveform based on **Equation 1a**. Frequency was derived as the reciprocal of the mean duration of time taken between each successive peak of the waveform based on **Equation 1b**, as suggested by Butt et al.^11^

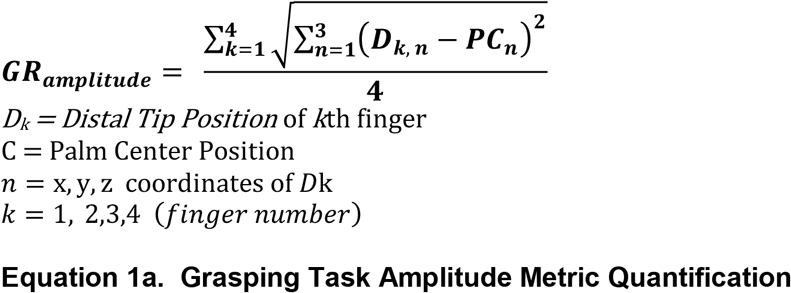

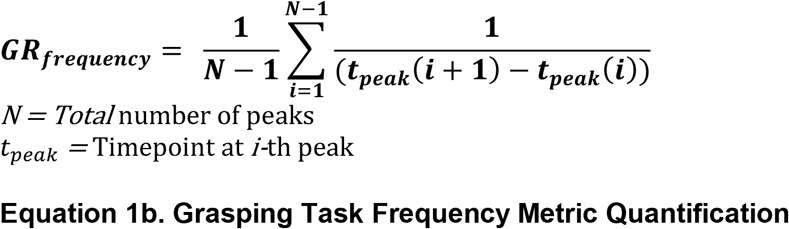

### 3. Study Design

Motor data collection using the LMC sensor was performed in two stimulation states: (1) DBS active using clinically effective, stable settings (DBS-ON) and (2) DBS turned off bilaterally (DBS-OFF) as noted in **Figure 1B**. The order of DBS-ON and DBS-OFF states were randomized such that the rater for motor movements was blinded to the DBS state. While most participants (73.3%) performed under on-medication, others (26.7%) were off-medication or reported unknown medication status. Because the entire testing session took only 26 minutes, the medication state was not observed to change during this time period. Patients continued on their normal medication regimen, allowing us to get a “real world” experience of motoric features that respond to acute DBS changes. Participants underwent a 10-minute wash-in or wash-out period in each stimulation state before testing to remove the residual stimulation effect from the previous DBS state before undergoing the motor test since this was previously shown to be sufficient to allow for the majority of motoric change from stimulation. ^14^

During the DBS-ON and DBS-OFF states, motor testing was conducted to evaluate bradykinesia by asking patients to perform the GR task of the MDS-UPDRS III. For each state, patients were asked to initially hold the movement for 3 seconds by looking at the “hold” grey cue on the computer screen, followed by 10 seconds of “go” green cue, followed by 3 seconds of “rest” grey cue. The “go” and “rest” cue sequence was repeated three times to collect three trials of motor data per hand (**Figure 1C)**. Patients were asked to perform movements starting with the right hand, followed by the left hand under each DBS state.

The severity of bradykinesia for each trial per hand was assessed using the MDS-UPDRS Part III scale by a trained, blinded rater, and the scores were averaged over the 3 trials for each stimulation state for each hand. In parallel, we simultaneously quantified amplitude and frequency motor features using the LMC sensor. The LMC sensor was placed directly underneath the palm to the maximize field of view of finger movements (**Figure 1D)**. A 3D-printed support apparatus was also used to allow study participants to rest their hands during the rest period of the task to maintain the constant height (∼30cm) between the hand and the sensor.

### 4. Lead Localization

For each patient who underwent LMC motor testing, a preoperative T1 MR and postoperative CT sequence were used for DBS electrode localization and volume of tissue activated (VAT) reconstruction using the Lead-DBS suite.^15^ First, the postoperative CT was co-registered to preoperative T1 and T2 MR sequences using the Advanced Normalization Tools (ANTs) + Subcortical Refine algorithm. Co-registered images then underwent normalization into a common MNI ICBM 2009b Nonlinear Atlas space using the ANTs. Bilateral DBS electrodes were reconstructed in a common atlas space from CT electrode trajectory artifacts using a refined TRAC/CORE method.^16^ After DBS electrode reconstruction, the VAT was generated based on activated contacts, stimulation parameters, and neighboring tissue conductivity. The spatial boundary of binary the VAT was defined as regions where the distribution of the electric field was 0.2 V/mm or higher as per previous investigations on neuron modeling. ^17-19^

### 5. Mean Effect Image (MEI)

A Mean Effect Image (MEI) was produced to display the spatial distribution of the degree of STN-DBS-induced motor feature change. For each patient, the DBS-induced percent change values (%) of amplitude and frequency of hand movements were assigned to voxels comprising the VAT on the contralateral hemisphere. These “numerical” VATs were then averaged across all patients in the cohort to produce a MEI based on previously published methods.^20, 21^ Regions with positive values in the MEI would suggest a cohort-level increase in that motor feature (frequency or amplitude) after DBS is applied, and in those with negative values are associated with a decrease in that motor feature.

### 6. Functional Connectivity and Motor Features

To identify cortical and subcortical regions that are significantly associated with post-DBS amplitude and frequency changes, we performed a correlation analysis between the functional connectivity maps produced by seeding a normative functional connectome with the VAT of each patient and respective percent change values (%) of each motor feature. This approach yielded an “R-Map”, a whole-brain map that represents the degree of correlation between functional connectivity strength of whole-brain regions with stimulation site and respective changes in motor features. For example, regions in R-Map with positive values notates region where the strength of functional connectivity is positively correlated with changes in motor features. This R-Map approach was adopted to assess how the degree of functional coactivation of neural activity between the subcortical stimulation site and cortical activity was associated with independent motor feature changes.

### 7. Statistical Analysis

Percent change values (%) of amplitude and frequency were correlated together for each hand using Spearman’s Rho correlation method with statistical significance set at p < 0.05. Furthermore, the change in amplitude and frequency were also correlated with the change in MDS-UPDRS III scores. All graphical and statistical analyses were performed using MATLAB 2021a (MathWorks, Natick, MA) and Prism 9 statistics package (GraphPad Software, San Diego, CA).

## Results

### 1. Demographics and Clinical Information

The demographics and clinical information of the study cohort are reported in **Table 1**. The study cohort consisted of 15 patients with STN-DBS, 13 of whom were males and 2 females. The majority of the cohort (86.7%) were right-handed. The reconstructed DBS electrodes for all 15 patients in the cohort are visualized in **Supplementary Material 1**.

**Table 1.** Demographics and Clinical Information. * Medication status of one patient was unknown.

| **Demographics** | ***N ± SEM (%)*** |
| --- | --- |
| **Sex** |  |
| Male | 13 (86.7%) |
|  | 70.4 ± 1.6 |
| **Age (yr)** |  |
| **Age at PD Diagnosis (yr)** | 57.2 ± 2.1 |
| **Duration of PD (yr)** | 13.2 ± 1.2 |
| **Handedness** |  |
| Right | 13 (86.7%) |
| Left | 2 (13.3%) |
| **Medication Status\*** |  |
| ON | 11 (73.3%) |
| OFF | 3 (20%) |
| **Most Affected Side\*** |  |
| Left | 7 (46.7%) |
| Right | 8 (53.3%) |

### 2. Association Between LMC Motor Features

Based on the LMC motor features during the GP task, the amplitude and the frequency were independently evaluated. The percent changes (%) of amplitude and frequency in the DBS-ON versus DBS-OFF states were then correlated against each other as shown in **Figure 2**. The change of frequency was negatively correlated with change in amplitude for both left (r = - 0.57, p = 0.035, **Figure 2A**) and right hand (r = -0.63, p = 0.014, **Figure 2B**).

**Figure 2:**
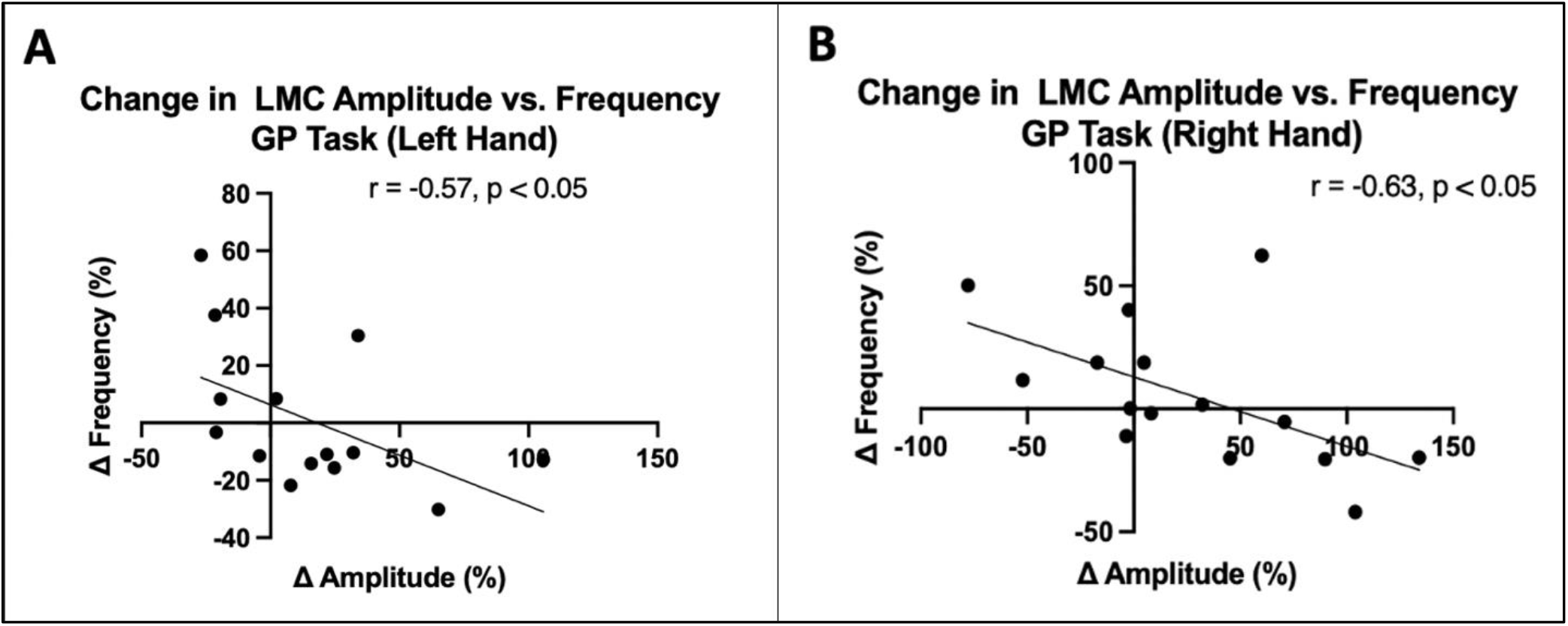
Association between LMC Amplitude and Frequency. (A) Left Hand. (B) Right hand. Correlation analysis was performed using Spearman’s Rank correlation. Both hands exhibited a negative correlation between the changes in amplitude and frequency after DBS was applied.

### 3. MDS-UPDRS III and LMC Motor Features

The strength of correlation was evaluated between changes in MDS-UPDRS III score during the GP task and each of the changes in LMC motor features - amplitude and frequency – caused by DBS activation. The correlation results in demonstrate that changes in MDS-UPDRS III features were only significantly correlated with the amplitude changes for both left (r = -0.63, p = 0.019) and right (r = -0.65, p = 0.01), but none with frequency changes (**Supplementary Material 2)**. This suggests that the assessment of DBS-induced changes in the MDS-UPDRS Part III hand grasp bradykinesia score is most sensitive to objectively measured changes in amplitude.

### 4. Mean Effect Image (MEI) of LMC Motor Features

By mapping the amplitude and frequency percent changes to the VAT for each patient, MEI of amplitude and frequency were produced. Based on both amplitude and frequency MEI, the changes in motor features are topographically represented across the stimulation sites in the STN. The amplitude MEI suggests a gradient of improved DBS-induced movement amplitude moving from anterior-medial regions to posterior-lateral regions (Figure 6). Conversely, stimulation across the same axis was associated with a decrease in frequency. This opposite trend is well reflected based on the spatial inversion of amplitude and frequency MEI in **Figure 3**.

**Figure 3.**
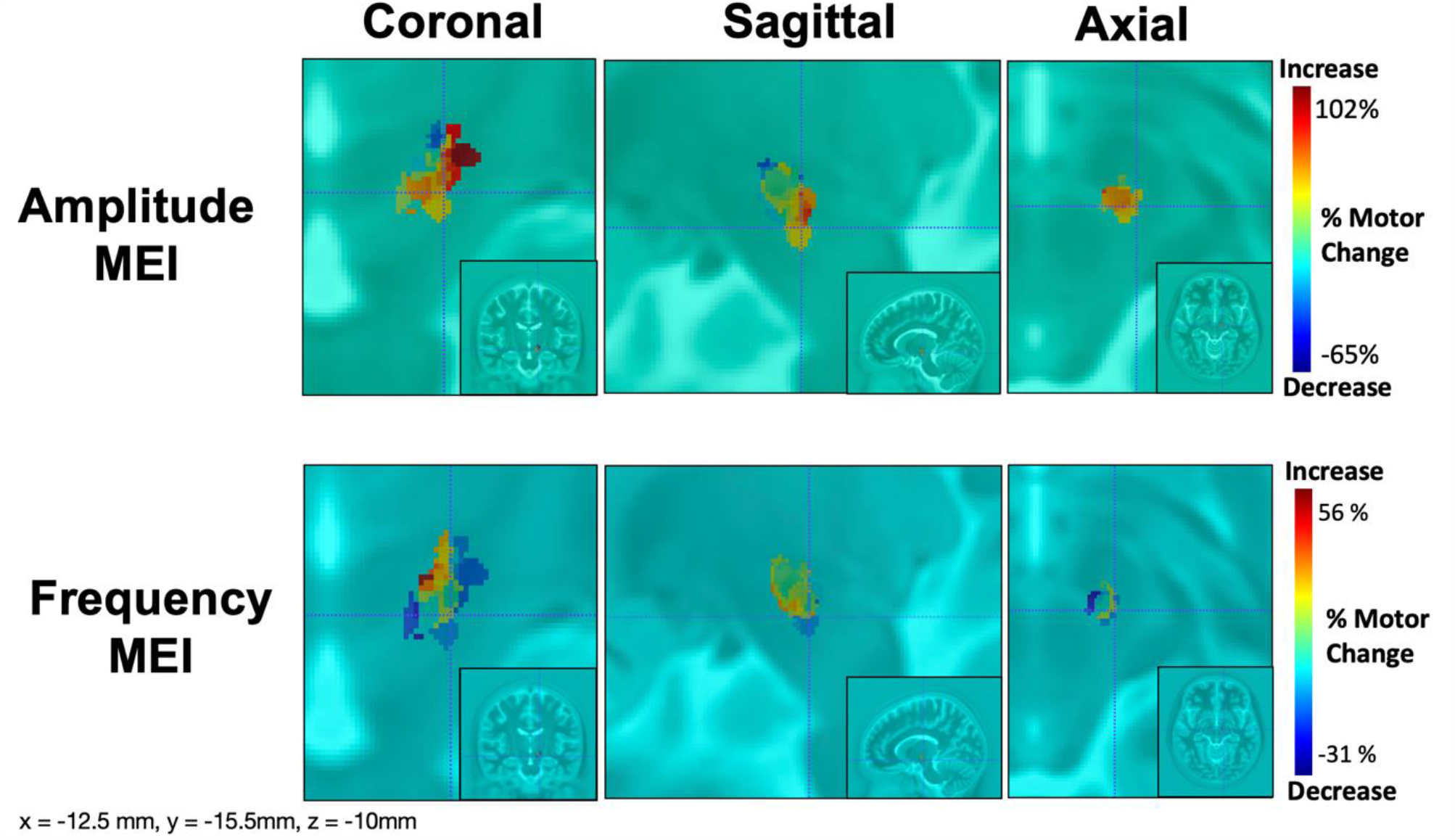
Mean Effect Image of LMC Motor Features. Amplitude and frequency MEI are visualized in the top and bottom row, respectively. The relative warmth or coolness of coloration represents correlation with an increase or decrease, respectively, in motor features relative to the mean effect.

### 5. R-Map of Amplitude and Frequency Changes

We observed a pronounced positive correlation between amplitude changes and functional connectivity strengths in prefrontal cortical regions (Figure 4). For right-hand GP tasks, larger amplitudes were associated with increased functional connectivity with prefrontal motor regions that included, but not limited to, left ventromedial prefrontal cortex (r = 0.579, p = 0.026), right middle frontal gyrus (r = 0.56, p = 0.03), and right medial superior frontal gyrus (r = 0.678, p = 0.007).

**Figure 4.**
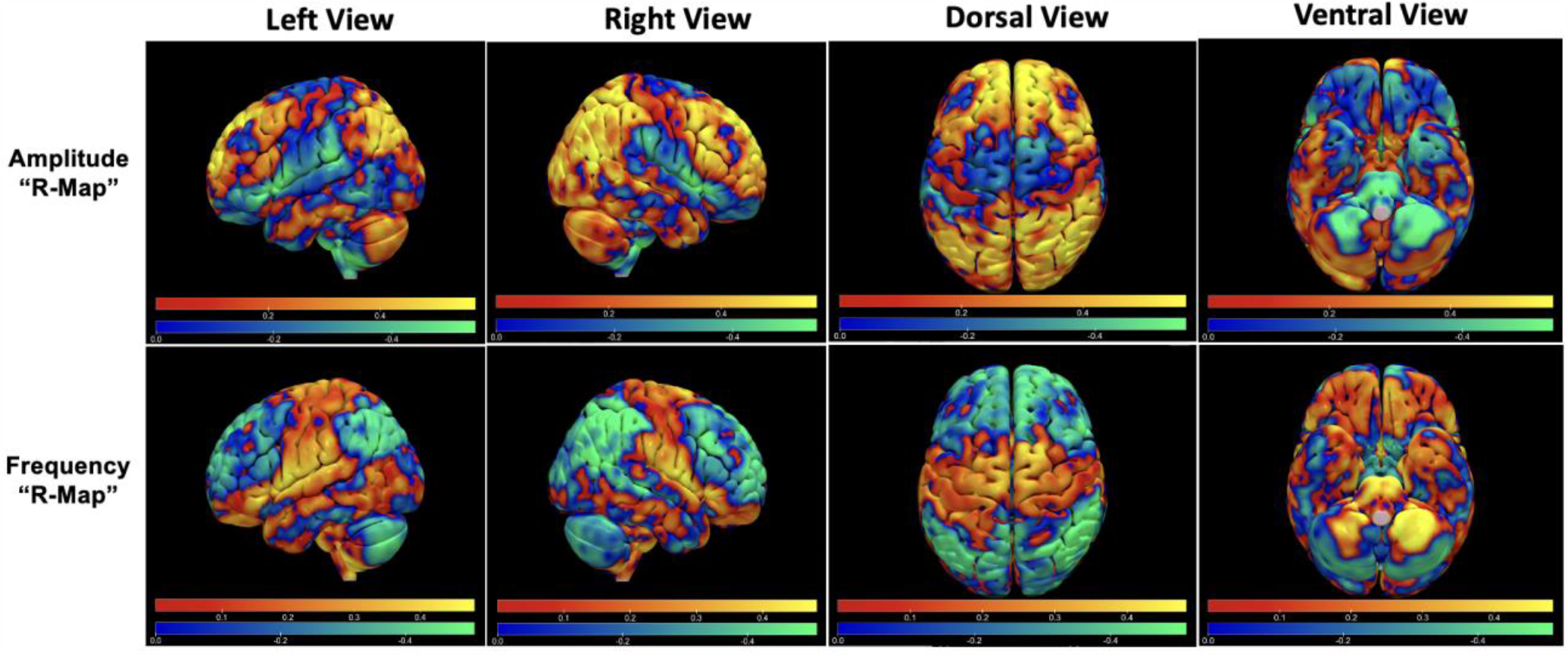
R-Maps of Amplitude and Frequency Changes Post-DBS. Red-Yellow regions represent areas with a positive correlation with changes in functional connectivity with the STN DBS VAT and LMC motor feature changes. Blue-Green regions represent areas with a negative correlation.

On the other hand, the R-Map for frequency changes was inverted, such that a negative correlation was observed between frequency changes and the strength of functional connectivity between the VAT and the prefrontal cortex. Increased frequency of grasping was associated with lower functional connectivity in the left ventromedial prefrontal cortex (r = -0.58, p = 0.025), right middle frontal gyrus (r = -0.58, p = 0.026), and right medial superior frontal gyrus (r = -0.57, p = 0.029).

## Discussion

In this study, we have investigated the distinctive effect of STN-DBS on components of bradykinesia - amplitude and frequency - in patients with PD. With the activation of clinically effective STN DBS, the changes in amplitude were negatively correlated with those in frequency, or vice versa. Furthermore, the DBS stimulation sites associated with an increase in amplitude were spatially distinct from those with an increase in frequency in the STN. This inversion of neural substrates representing amplitude and frequency changes persisted from the subcortical basal ganglia (BG) level to the global cortical level through whole-brain connectivity analysis. To the best of our knowledge, this study is the first to report the anatomical segregation of the STN motor sub-regions that differentially encode amplitude and frequency changes post-DBS, and how this segregation was maintained across functional connectivity patterns at the cortical level.

We have closely considered the relationship between amplitude and frequency in the grasping task and have demonstrated a significant negative correlation between amplitude and frequency changes observed in both hands (r = -0.57 for left hand, r = -0.63 for right hand) after DBS; as amplitude increases, the mean frequency slows. If velocity is relatively constant, this trade-off would be inherent to increasing amplitude. However, this negative correlation is not solely explained by this trade-off with reduced frequency as amplitude increases since there were stimulation settings that improved frequency at the cost of amplitude. (**Figure 2**). Interestingly, the association between these motor features and the MDS-UPDRS III bradykinesia rating further revealed the changes in bradykinesia were significantly driven by changes in amplitude, rather than frequency. Specifically, the increase in amplitude was correlated with the improvement of bradykinesia scores for both hands despite the slowing of hand grasping with larger amplitudes. While we found no previous studies have explored the direct effect of DBS on bradykinesia components, one study reported that MDS-UPDRS Part III was also negatively correlated with amplitude during GP task using a wearable kinematics device in the baseline condition.^22^

After the changes in motor features were mapped to volumes of stimulated tissue across all subjects to create the MEI, the MEI valence (“sweet spot” versus “sour spot”) was inverted between amplitude and frequency changes: stimulation location with the *maximum increment* of amplitude corresponded to that with the *maximum* decrement of frequency. The “sweet spot” for amplitude changes was more posterior-dorsolateral and for frequency, more anterior-medial in the STN. One could hypothesize that “effective” STN stimulation is increasing amplitude but decreasing frequency and that the frequency “sweet spot” is just the absence of a robust amplitude benefit. However, participants with stimulation of this region actually shows improved frequency over DBS-Off, at the cost of amplitude (smaller, faster movements compared to DBS-OFF).

Based on this spatially graded distribution within STN, we postulate that the negative correlation between amplitude and frequency changes was driven by the inversive representation within the shared neural substrate and the network connectivity of these two different STN subspace. R-maps produced between amplitude and frequency were spatially inverted, with regions with significant correlations in the middle frontal gyrus, medial superior frontal gyrus, and ventromedial prefrontal cortex. In a sequential movement such as the GP task, the pre-supplementary motor area (pre-SMA), part of the medial superior frontal gyrus and medial frontal cortex, has been suggested to be involved in the initiation of self-generated action sequences.^23-26^ During a self-initiated movement task, the functional connectivity between STN and pre-SMA region has been reported to be strengthened for both healthy and PD patients by employing the hyperdirect and indirect basal ganglia pathways.^27^ STN-DBS may interfere in the STN-pre-SMA connection to modulate performances in movement initiation in GP, which may emerge as alterations in the motor frequency and amplitude from baseline condition. The magnitude and the direction changes of two motor features (i.e increment or decrement) after STN-DBS may be determined based on the locations of individual VATs relative to “amplitude hotspots” or “frequency hotspots” spatially distributed across the dorsolateral-ventromedial axis of the STN. However, the precise STN-pre-SMA connections associated with amplitude and frequency changes are yet unknown and would warrant further investigations.

The spatial distribution of DBS’s differential effect on amplitude of rapid movements are consistent with existing literature. That targeting dorsolateral STN for DBS was associated with the greatest bradykinesia improvement ^6, 28^ is consistent with our finding, such that posterior dorsolateral STN resulted in the greatest improvement of amplitude changes and the greatest improvement in MDS-UPDRS III bradykinesia rating. The dorsolateral region of STN has further been shown to share strong structural connectivity with sensorimotor motor cortical areas such as the primary motor cortex and supplementary motor areas (SMA). Conversely, the more medial region of STN encompassing the “sweet spot” for frequency increment, is spatially proximal to the associative subregion of STN that shares strong connectivity with the associative areas of prefrontal cortices. ^29, 30^ The differential responses in amplitude and frequency changes thus may be attributed to the activation of separate STN-prefrontal networks from DBS. Expanding on the network-level stimulation effect on motor features, we have performed an R-map analysis correlating the degree of functional connectivity seeding VATs and changes in motor features.

Several limitations exist in this study. This is a preliminary study performed across a limited cohort of 15, and future studies with a larger sample size are strongly encouraged to replicate our findings. Furthermore, in addition to amplitude or frequency, the MDS-UPDRS-III rating is composed of other motor features, including hesitations or temporal decrement. We chose to focus on amplitude and frequency because of their canonical influence on shaping the bradykinesia rating, and clear pre-processing steps to evaluate them from raw motor data. Future studies should aim at how performances in self-generated action sequences as GP-task are reflected and altered across multiple motor features of bradykinesia after STN-DBS. Lastly, we focused on the isolated effect of STN DBS and aimed to run our 26-minute paradigm during whatever medication state in which they presented, hoping to look at the subtle changes in bradykinesia detected with the LMC even in the fully or partially medicated state. It is possible that medication status changed during our paradigm. Nevertheless, because the paradigm was relatively short, we are confident that both DBS-ON and DBS-OFF test periods were performed in the same state of medication effect. Regardless, we randomized the stimulation state (“on” or “off”) so that if a medication wearing-off or kick-in effect were to occur, it would not systematically be linked with the stimulation “on” or “off” state.

## Conclusion

In this study, we demonstrated a differential effect of STN-DBS on bradykinesia motor features (amplitude and frequency), an effect that also translates to differential network engagement. Improvement in hand movement amplitude were negatively correlated movement frequency, and were associated with stimulation of a different STN subregion than that which was associated with an improvement in frequency and a decline in amplitude. The degree of motor changes was represented as a spatial gradient across the ventromedial–dorsolateral axis of the STN. Functional connectivity analysis between stimulated volumes in STN and cortical areas showed a differential pattern for improvement in amplitude versus frequency of movements.

## Data Availability

All data produced in the present study are available upon reasonable request to the authors

## Author Roles

1. Research project: A. Conception, B. Organization, C. Execution;
2. Statistical Analysis: A. Design, B. Execution, C. Review and Critique;
3. Manuscript Preparation: A. Writing of the first draft, B. Review and Critique;

M.J.K.: 1A-C, 2A-B, 3A

Y.S.: 2A, 2C, 3B

J.L.: 1C

Y.S.: 1A, 3B

W.S.A.: 1A, 3B

K.A.M.: 2C, 3B

---

**Supplementary Material 1:**
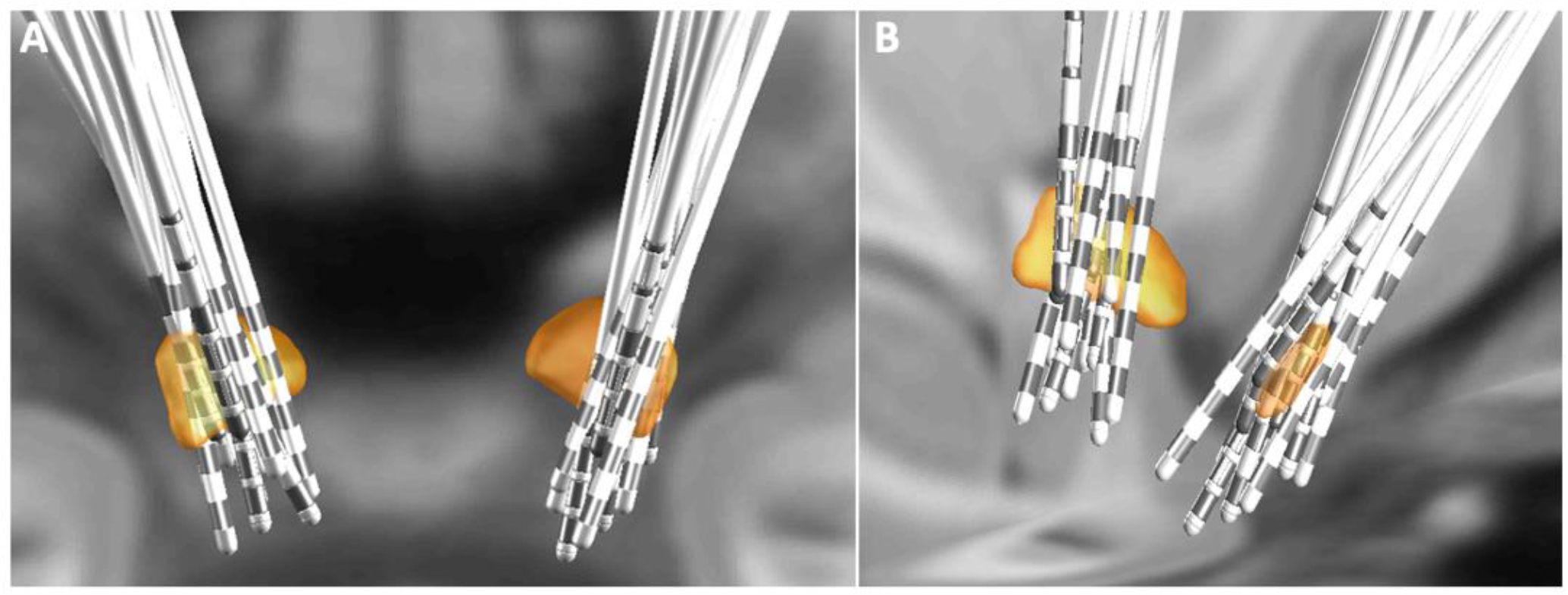
Grouped STN-DBS Lead Localization. DBS leads of 15 patients are reconstructed and shown with bilateral STN (orange) in **(A)** dorsal and **(B)** parasagittal view.

**Supplementary Material 2:**
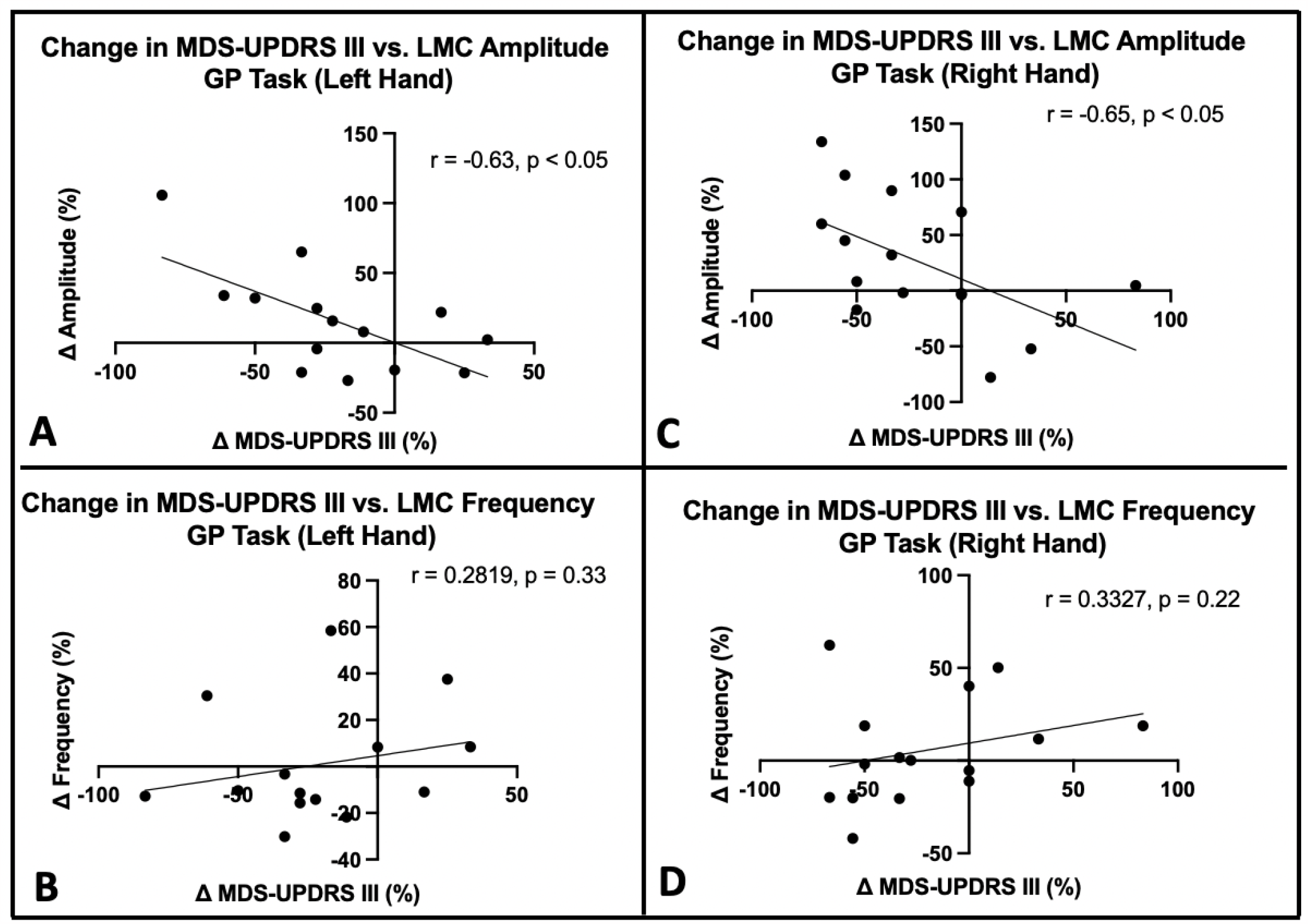
Association between LMC Motor Feature Changes and MDS-UPDRS III Rating Changes. Correlation of changes in MDS-UPDRS III with change in amplitude (top row) or frequency (bottom row) in left hand (left column) or right hand (right column). Correlation coefficients shown for statistically significant correlations.

